# Prediction of dynamic balance state and recovery following stroke using fMRI graph analysis

**DOI:** 10.1101/2024.10.28.24316247

**Authors:** Or Symonitz, Katherin Joubran, Rami Puzis, Lior Shmuelof

## Abstract

Stroke is associated with damage to neural tissue and is the leading cause of long-term sensory-motor disability in adults. Dynamic balance impairments are one of the most debilitating outcomes of stroke, leading to increased falls and loss of mobility. While the recovery of motor functions following stroke was shown to be affected by the initial brain damage, the ability to predict recovery based on neural markers is limited due to the involvement of multiple brain areas in dynamic balance, and the limited size of available datasets. We apply graph-theory-based neural markers to predict the extent of recovery in the presence of rehabilitative treatment and the passage of time on a dataset of 21 subjects after stroke. We report that global features are more informative than local features, describing individual regions. We also report that recovery level is predicted more accurately (85%) than dynamic balance state (76%). Our results demonstrate the feasibility of graph-based analyses on limited datasets and may contribute to clinical goal setting and to mapping the neural substrates of dynamic balance.

## 1 Introduction

Ischemic stroke due to a disruption of the blood supply is characterized by initial neuronal loss and physiological and chemical neural changes (i.e., focal/connectional diaschisis) (Tang et al. 2015, C. Tang et al. 2016, Carrera and Tononi 2014). These changes, which are caused by the primary brain lesion or by the modulation of neuronal networks far from the brain lesion (Carrera and Tononi 2014), cause physical and functional impairments (Nudo 2013, Hylin, et al. 2017). Mobility impairments, such as dynamic balance deficits, are a prominent outcome of brain injury (McCulloch et al. 2010), with 73% of stroke survivors reporting impairments in dynamic balance control, and an increased risk of falls during the first six months post-stroke (Forster and Young 1995).

Dynamic balance control is a multifactorial ability that depends on feedback and feedforward control mechanisms and is mediated by both central pattern generators at the spinal level and subcortical and cortical brain areas (Rossignol et al. 2006). Partial recovery of dynamic balance impairments following injury occurs due to the passage of time and the rehabilitation treatment. However, there is high inter-subject variability of recovery which can be partially explained using neural measures, such as lesion size and corticospinal tract integrity (Stinear et al. 2014).

Functional magnetic resonance imaging (fMRI) can be used for estimating neural connectivity changes following stroke. Specifically, resting state fMRI (rs-fMRI) is a popular methodology for estimating functional connectivity due to its non-invasive nature, good agreement with task-related connectivity (Biswal et al. 1995, Cordes et al. 2000, De Luca et al. 2005), and the lack of movement confounds. Indeed, Almeida et al. (2017) reported that stroke subjects with motor impairments had higher resting state functional connectivity (FC) between the primary motor cortex (M1) and the contralesional hemisphere than healthy control subjects. We recently showed that response to treatment is associated with reduced FC in the sensorimotor and cerebellar networks (Joubran, Bar-him 2022). However, the high dimensionality of the rs-fMRI connectivity data, the limited size of clinical datasets, and the involvement of multiple neural markers in dynamic balance (Takakusaki, 2017), limits the replicability of these measures and their future applicative contribution.

One way to reduce the dimensionality of the data is by extracting meaningful global and local measures from the rs-fMRI correlation network. Such network-based measures were shown to be useful for classification tasks related to brain impairments (Khazaee et al. 2016, Jiang et al. 2023, Du et al. 2024). In this study, we aim to predict the dynamic balance state and recovery in stroke patients by using graph-based measures in combination with machine learning algorithms. A major challenge in machine learning research of such a specific problem is the absence of large, labeled datasets. Provided with a small dataset, we identify robust global features that predict an improvement in dynamic balance (Community Balance and Mobility scale (CBM)) following rehabilitation treatment. This paper demonstrates how to identify informative features and ensure their robustness despite the limited data. Finally, we discuss the mechanisms involved in dynamic balance impairments and recovery considering our findings.

## 2 Methods

### 2.1 Summary of methods

A group of twenty-one participants with stroke participated in this study. Resting state functional magnetic resonance imaging data (rs-fMRI) and community balance and mobility scale (CBM) measures were taken before (T1) and after (T2) a three-months rehabilitation period. 13 subjects at T1 and 10 at T2 had low CBM scores (<39) (Balasubramanian 2015). After the rehabilitation period, 10 subjects recovered CBM (Δ*CBM = CBM*_*T2*_ *- CMB*_*T1*_) by more than 5 points, suggesting recovery.

Three binary prediction tasks are considered in this study:

- **P1**: **baseline dynamic balance ability:** Predicting a high/low CBM score from the patient’s pre-treatment scan (T1).
- **P2: post-treatment dynamic balance ability:** Predicting a high/low CBM score from the patient’s post-treatments scan (T2).
- **P3: dynamic balance recovery**. Predicting a high/low CBM recovery using the patient’s pre-treatment scan.

All prediction tasks are performed using the same machine learning pipeline comprising nine stages (Figure 1). For each prediction task, we integrated global features, local features, embedding coordinates, and edge filtering methods of the functional brain graphs constructed from the rs-fMRI scans. Due to the small size of the dataset, the entire pipeline (feature selection, hyperparameter tuning, and training) was evaluated according to the nested leave-one-out cross-validation procedure (NLOOCV), see Section 5.9). In addition, to address the concern of leakage of information from test data into the trained models, we describe the robustness of selected features and model parameters under perturbations. We consider the following performance indicators to assess the quality of the predictors:

**Figure 1:**
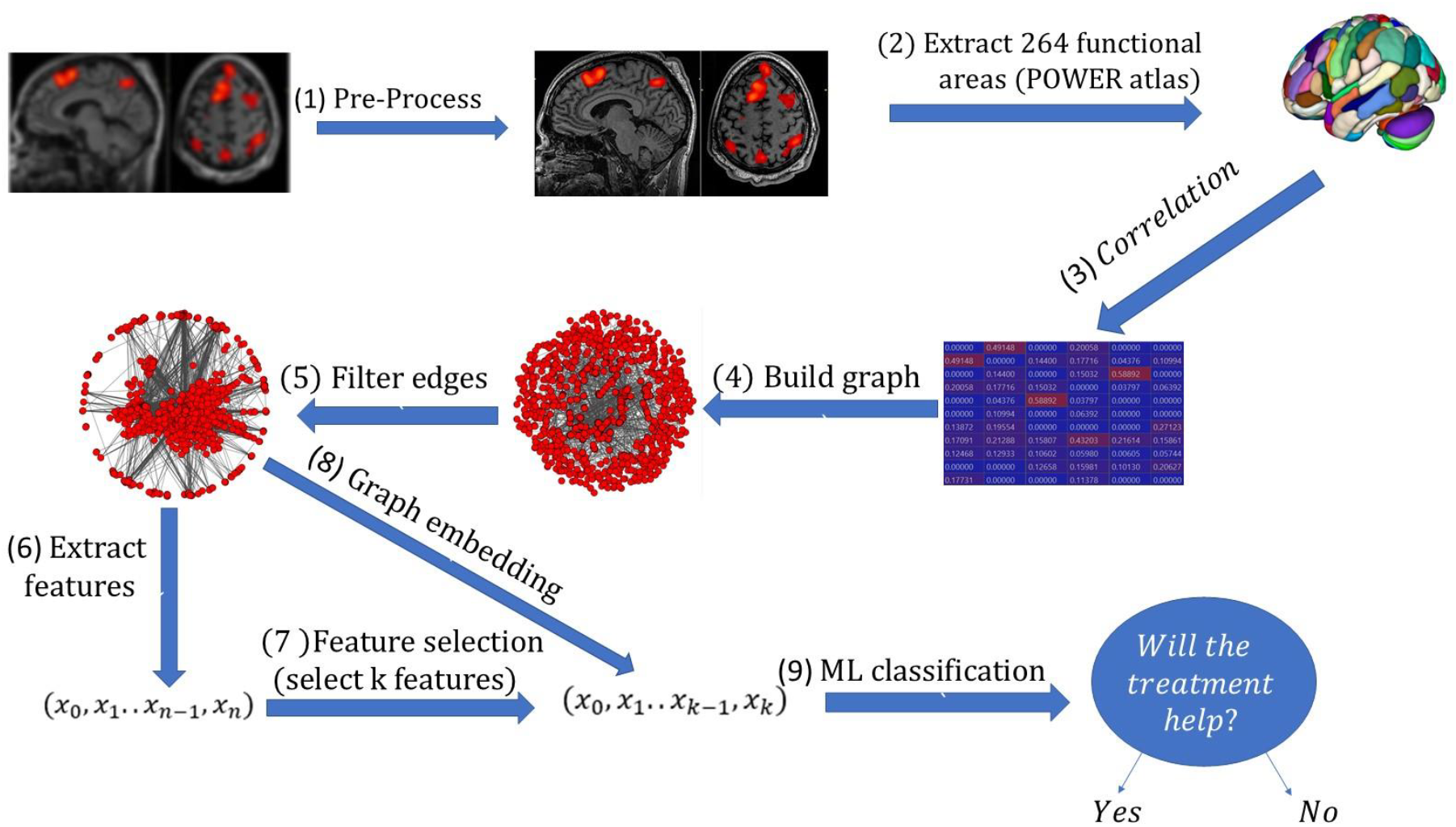
The analytical pipeline used in this research includes 9 steps: (1) preprocessing the data, (2) extracting functional areas from the rs-fMRI, (3) computing a correlation matrix from the 264 ROIs, (4) building the FC graph (ROIs are nodes, and edges are derived from correlations), (5) filtering edges using different criteria, (6) extracting features (local or global features) (7) selecting the features. Stages 6 and 7 can be replaced with stage 8 (graph embedding). The last step is (9) using the features for the classification task.

- Prediction accuracy (acc), measured as the proportion of correct predictions.
- Parameter sensitivity, defined as the range of the optimal parameter values.
- Feature selection consistency, measured as the number of times a feature was selected.

### 2.2 Data

#### Subjects

Twenty-one stroke subjects (14 males and seven females), at least six months post-stroke, participated in the study, which was approved by the Institutional Reviewing Board. The age range of participants was 47-80 (years), with an average age of 65.81±7.33 (mean ± std).

The dynamic balance rehabilitation was performed at Reuth Rehabilitation Medical Center in Tel Aviv, Israel. Clinical examinations were conducted at Ben-Gurion University of the Negev in Beer-Sheva. FMRI scans were performed at Soroka University Medical Center in Beer-Sheva. See the ethics statement at the end of this article.

#### Experimental procedure

Each subject was assessed twice; before (T1) and after (T2) receiving rehabilitation treatment. The treatment consisted of 22 sessions administered over a period of nearly three months. Both assessments included (1) a clinical assessment in the lab using the Community Balance and Mobility Scale (CBM) and (2) an rs-fMRI scan. The CBM values were 33.71±15.18 (mean ± std) at T1 and 38.23±15.74 (mean ± std) at T2. The effectiveness of the treatment was assessed by computing the difference between T1 and T2 CBM (ΔCBM), and was found to be 4.52±6.64 (mean ± std) (Joubran et al., 2021)

#### Imaging protocol

The imaging was performed at Soroka University Medical Center’s imaging center using a 3-Tesla Philips Ingenia whole-body MRI scanner. Two similar imaging sessions were conducted, before (T1) and after (T2) rehabilitation treatment. The rs-fMRI data was acquired using a gradient echo EPI with a voxel size of 3×3×3 millimeters (mm), TR=2000 milliseconds (ms), TE=35ms, flip angle=77°, 35 slices, with a 0.6 mm gap. The first two volumes were discarded to allow magnetization to reach equilibrium. The length of the resting state scans was seven minutes and 26 seconds.

### 2.3 Pre-processing

Pre-processing of the rs-fMRI data included removal of the first two functional images of each run series to allow stabilization of the magnet; slice time acquisition correction (ascending-interleaved; using a cubic-spline interpolation algorithm); head motion correction (trilinear interpolation); and temporal filtering: a high-pass (GLM-Fourier) frequency filter with a cut-off value of two sine/cosine cycles and a low-pass Gaussian full width at half maximum (FWHM) of 1.9 data points to remove unwanted physiological signals such as respiratory and heart rate (Fox et al., 2007).

Further, we used band-pass filtering with an eighth order Butterworth filter with cut-off frequencies of 0.009Hz and 0.08Hz (Fox et al., 2007). Data was coregistered to the anatomical scan and transformed to the Talairach space (Talairach, 1988). Pre-processing was conducted in BrainVoyager 20.6 and was similar to the pre-processing in Joubran et al., 2022.

### 2.4 Prediction tasks

Three binary prediction tasks were considered:

P1. Predicting the high/low CBM score (T1) from the patient’s pre-treatment scan..

P2. Predicting the high/low CBM score (T2) from the patient’s post-treatments can (T2).

P3. Predicting the high/low CBM recovery (T2-T1) using the patient’s pre-treatment scan denoted in this paper as Δ*t*_*CBM*_.

To ensure that the proposed classifiers rely on non-trivial constructs we used the following baseline prediction methods:

1. Zero-rule – a constant prediction across subjects that always predicts the majority class.
2. CBM-based classifier – relies only on the T1 CBM score to predict Δ*CBM* in the P3 problem. [this is the only place that this term is used. Add it to the relevant analysis in results]

### 2.5 Graph construction

We constructed a functional brain graph *G* = (*V, E*) for each subject based on the rs-fMRI scans. The nodes (V) correspond to 264 pre-defined regions of interest defined by the POWER atlas (Dosenbach et al. 2010). The edges *E* are derived from the absolute Pearson’s correlation coefficient between the nodes’ activation vectors. We removed edges with negative correlations. Three threshold filtering criteria were used to binarize the correlation coefficients:

1. **Fixed threshold.** Edges with a Pearson correlation coefficient above a predefined threshold are retained. For each examined threshold *t*, the same threshold *t* was used for all subjects.
2. **Density threshold.** By weighting each edge with a Pearson correlation as it weight, the *m*% heaviest edges from each graph (for each subject) are retained. This method allows us to control for the number of edges in each graph.
3. **Planar graph.** Planar graphs are constructed by eliminating edges with low correlation coefficients, using the PMFG (planar maximally filtered graph) method (Tumminello et al. 2005). The algorithm uses the efficient computation of the planar graph which was suggested in (Boyer and Myrvold 2004). This method is commonly used in conjunction with correlation-based graph analysis (Song et al. 2011).

### 2.6 Expert-based feature extraction

To apply machine learning techniques on functional connectivity (FC) graphs for classification tasks, one needs to first extract or generate features that capture essential aspects of the graphs. Such features can be defined based on prior knowledge.

One way to categorize network features is by (1) local features describing specific nodes or edges and (2) global features describing an entire graph. The degree of connectivity (the number of edges connected to a node) and the local clustering coefficient (the fraction of triangles a node participates in out of all pairs of its neighbors) are examples of local features (the full list can be found in supplementary materials). Since the set of nodes in an FC graph is fixed when relying on a specific atlas, feature vectors of all nodes can be concatenated to produce a feature vector of a graph.

Graph theory provides a variety of global features, including density, algebraic connectivity, diameter, number of connected components, transitivity, and rich-club coefficient, to name a few Bondy, Murty et al. (1976) (Full list can be seen at supplementary material). In addition, global features can be produced by aggregating local features, e.g. average degree, the kurtosis of local clustering coefficient. The four aggregations used in this study are averaging, variance, skewness and kurtosis.

### 2.7 Feature selection

A set of 587 global and aggregated local features were extracted, in addition to 30 to 264 rich-club coefficients (depending on the density of the graph). Due to the large number of features and the consideqrably small dataset, we selected a small number of features to prevent overfitting. Feature selection was performed based on information gain ratio (Quinlan 1986) between features in the training set and the target variables (high/low CBM, high/low Δ*CBM*). Information gain quantifies the dependence between a random variable and the target variable. It indicates the usefulness of a feature for predicting the target. We computed information gain using the scikit-learn package. We chose the *k*∈ [1, 9] features with the highest score.

### 2.8 Graph embedding

Expert-based feature extraction is not the only method for representing graphs as fixed size numeric vectors. Graph embedding algorithms derive vector representations of graphs based on some graph similarity measure. In this study we used Family of Graph Spectral Distances (FGSD) (Verma and Zhang 2017), graph2vec (Narayanan et al. 2017), Spectral Features embedding (de Lara and Pineau 2018), Graph and Line graph to vector (GL2VEC) (Chen and Koga 2019) and two variants of Network Laplacian Spectral Descriptor (NetLSD) (Tsitsulin et al., 2018). NetLSD is based on spectral graph theory; this method uses graph signatures based on heat (NetLSD-heat) or wave (NetLSD-wave) kernels of the Lapalacian which inherit the formal properties of the Laplacian spectrum. All graph embedding algorithms were examined under the same experimental setup. Unsupervised graph embedding algorithms produce generic features that are good for many tasks, including classification, without the need for specific domain knowledge. Good unsupervised graph embedding encodes the main differences between the graph topologies in a short vector. For example, two graphs that have similar global features should be found in vicinity to one another in the embedding space. Similar to expert-based features, we use only few dimensions (d ∈[1, 9]) for graph embedding to avoid overfitting due to the small amount of data. Note that the number of dimensions is a common parameter of the graph embedding algorithms and feature selection is not required.

### 2.9 Supervised machine learning

We framed the three prediction problems P1, P2, and P3 as binary classification tasks. The target binary variable in P1 and P2 is low/high CBM, as defined by a threshold *t*_*CBM*_ = 39. The target binary variable in P3 is low/high Δ*CBM*, as defined by a threshold *t*Δ_*CBM*_ = 6. The thresholds were chosen as the median CBM and Δ*CBM* values to create balanced datasets. The values *t*Δ_*CBM*_ = 6 and *t*_*CBM*_ = 39 were validated by experts as reasonable clinical thresholds.

After extracting and selecting the features, we used a random forest (RF) classifier to solve the classification tasks P1, P2, and P3. RF is an ensemble model relying on multiple decision trees.

### 2.10 Nested cross-validation

For each prediction problem we split the dataset into three distinctive sets – train set which we would using for training our model, validation set which be used for optimizing parameters and a test set for testing our model. Due to the small amount of data, validation and hyper-parameter tuning are performed using the nested leave-one-out cross validation procedure (NLOOCV). Each of the 21 subjects is used to test models that were trained on the other 20 subjects. LOOCV allows utilizing all the data for testing while preventing leakage of ground-truth information into the models and assessing the robustness of the models on multiple perturbed training sets.

The FC graph construction, feature extraction and generation, and supervised machine learning methods used in each iteration of the LOOCV are all parameterized. These hyperparameters should be optimized to obtain favorable performance. Machine learning best practice suggests splitting the data into training, validation, and test sets to fit the models, optimize hyperparameters, and evaluate the resulting models respectively.

Due to the limited size of the dataset, we applied LOOCV for hyperparameter optimization (validation loop) in each iteration of the testing LOOCV (testing loop). This process is known as nested LOOCV (Cawley and Talbot 2010). In every iteration of the testing loop, the models are trained 20 times on 19 subjects and are validated on the remaining subject to choose the best set of hyperparameters. Then, after choosing the hyperparameters, we train a model on all the 20 subjects and apply it on the remaining test subject.

Different parameters were selected for each test-fold based on the inner LOO tuning process. Then, we trained a new model based on all the data without the test set, using the selected parameters and later predicted the test subject. We examined the effects of the number of selected features and the different thresholds. As mentioned earlier, we used *k* ∈ [1, 9] for the number of features and for the embedding dimension. We examined the same numbers for both the feature selection and the embedding dimension, since they are both used as the number of dimensions for the model. As mentioned, we only examined one to nine dimensions.

The entire nested cross-validation described above is presented in Figure 2. There is no information leakage during this procedure. Both the stages of feature selection and the model training did not use the test data.

**Figure 2:**
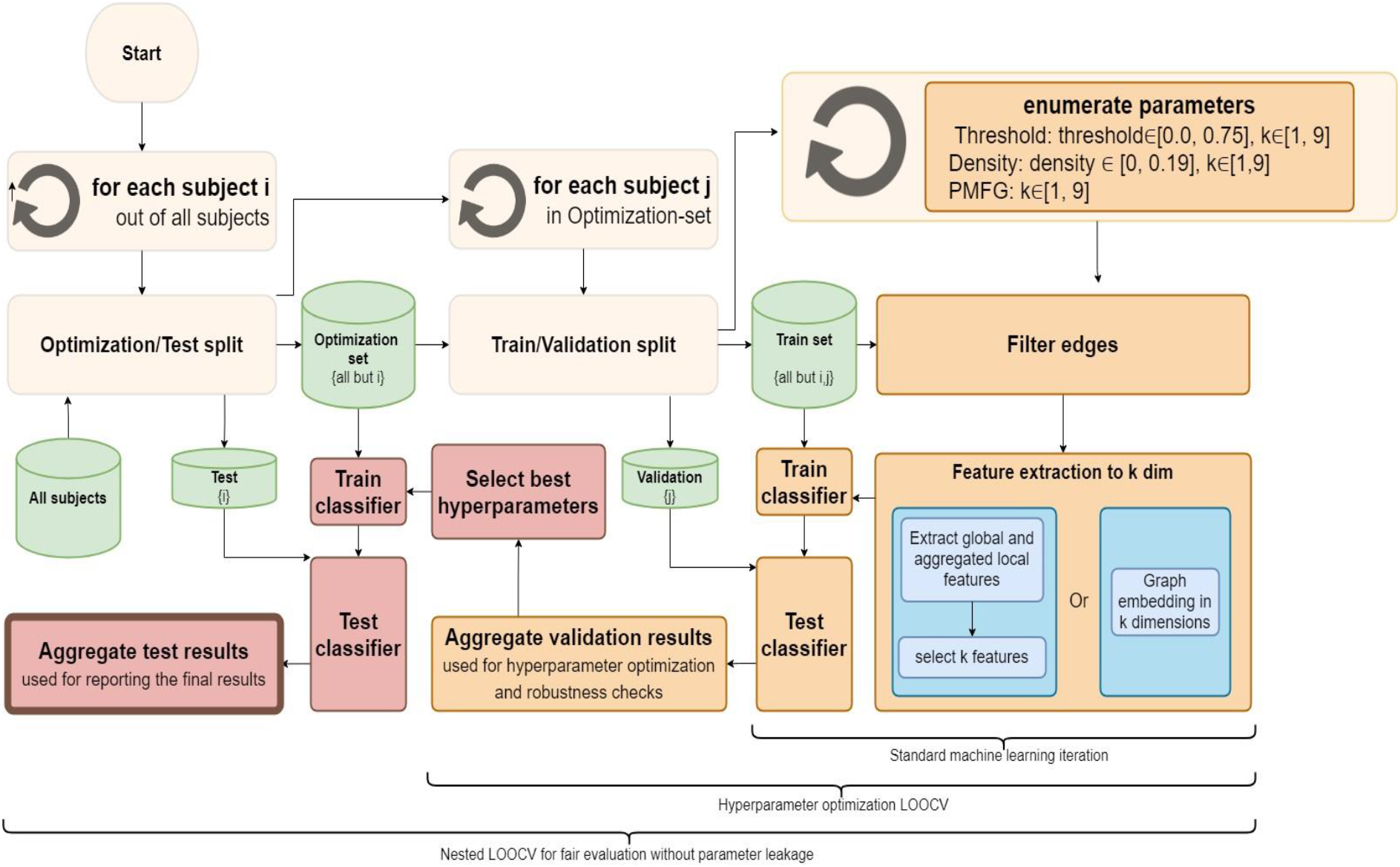
Hyperparameter optimization using the nested leave-one-out method..

### 2.11 Measuring robustness of the results, features, and hyperparameters

Given the small dataset, overfitting or learning from noise is a serious concern. To address this concern, we measured the robustness of the results, hyperparameters and features. Robustness of the results was defined as constant performance across consecutive values of hyperparameters. The variance of those results should be as small as possible.

Robustness of the hyperparameters is defined as the probability of selecting the same hyperparameters in the nested LOOCV. The variance of the hyperparameter list is a measure of this robustness. Robustness of the features is defined as the probability of selecting the same features in the nested LOOCV, which is quantified as the total number of times each feature was selected.

## 3 Results

### 3.1 CBM prediction from respective scans

We examined the accuracy of classifying subjects to high (*>*= 39) or low (*<* 39) CBM scores at T1 and T2, given the respective rs-fMRI scans of the 21 subjects (tasks P1 and P2 respectively). This analysis highlights the relation between the structure of a functional brain network and CBM.

#### Prediction accuracy

The best accuracy for T2, (0.76), was obtained using the fixed threshold criteria for edge filtering (see Section 3.6) and NetLSD-heat graph embedding (see Section 3.9). PMFG criteria received an accuracy score of 0.76 with 5-9 features. The results, summarized in Table 1, show that the best predictors perform better on T2 data than on T1 data. The edge filtering thresholds selected during hyperparameter tuning fall within the range (0.4-0.69) regardless of the chosen feature sets and scans (T1/T2), and the best edge filtering densities fall within the range (0.01-0.19).

**Table 1.** Results of the tasks with the different edge criteria, feature sets and number of features for predicting high/low CBM after rehabilitation treatment.

| Edge filtering criteria | Feature set | Avg. P1 test acc. | Selected thresholds (min-max) | Num. of features (min-max) | Avg. P2 test acc. | Selected thresholds (min-max) | Num. of features (min-max) |
| --- | --- | --- | --- | --- | --- | --- | --- |
| Zero-rule |  | 0.62 |  |  | 0.52 |  |  |
| Threshold | Expert-based | 0.57 | (0.42, 0.68) | (1, 9) | 0.66 | (0.4, 0.43) | (1,9) |
|  | NetLSD-heat | 0.66 | (0.44, 0.46) | (3,6) | **0.76** | (0.4, 0.44) | (2, 9) |
|  | NetLSD-wave | 0.52 | (0.42, 0.64) | (2, 8) | 0.52 | (0.4, 0.6) | (2, 9) |
|  | Graph2Vec | 0.33 | (0.4, 0.66) | (1,5) | **0.76** | (0.43, 0.69) | (1, 9) |
|  | FGSD | 0.71 | (0.45, 0.64) | (1, 3) | 0.57 | (0.41, 0.44) | (1, 4) |
| Density | Expert- based | 0.38 | (0.02, 0.18) | (1, 9) | 0.47 | (0.03, 0.16) | (1, 9) |
|  | NetLSD-heat | 0.66 | (0.01, 0.05) | (2, 9) | 0.43 | (0.04, 0.16) | (1, 9) |
|  | NetLSD-wave | 0.38 | (0.04, 0.12) | (2, 9) | 0.24 | (0.04, 0.19) | (2, 9) |
|  | Graph2Vec | 0.43 | (0.01, 0.18) | (1,9) | 0.62 | (0.04, 0.13) | (1, 9) |
|  | FGSD | 0.33 | (0.05, 0.12) | (1, 8) | 0.095 | (0.02, 0.16) | (9, 1) |
| PMFG | Expert-based | 0.24 |  | (1, 9) | 0.71 |  | (1, 9) |
|  | NetLSD-heat | 0.62 |  | (1, 9) | **0.76** |  | (3, 9) |
|  | NetLSD-wave | 0.71 |  | (3,7) | 0.19 |  | (1, 8) |
|  | Graph2Vec | 0.57 |  | (1, 9) | 0.38 |  | (1, 9) |
|  | FGSD | 0.33 |  | (1, 9) | 0.57 |  | (2, 4) |

### 3.2 Predicting CBM recovery using pre-treatment scan

Here we report the performance of predicting the high/low CBM recovery after rehabilitation treatment (P3). This is the main task evaluated in current study.

#### Prediction accuracy

The best test accuracy (0.857) was obtained using the threshold filtering criteria and global graph features (see Table 2). Since the Δ*CBM* dataset is balanced (the number of subjects with high/low Δ*CBM* is 10/11 respectively), the zero-rule baseline’s accuracy is 0.52. FGSD embedding with one dimension and the density filtering criteria of 0.02 performed better than zero-rule, achieving an accuracy of up to 0.76. The results reported here were obtained with different hyperparameter values and selected features, each chosen during the respective validation iteration. The ranges of the hyper-parameter values are presented in Table 2.

**Table 2:**
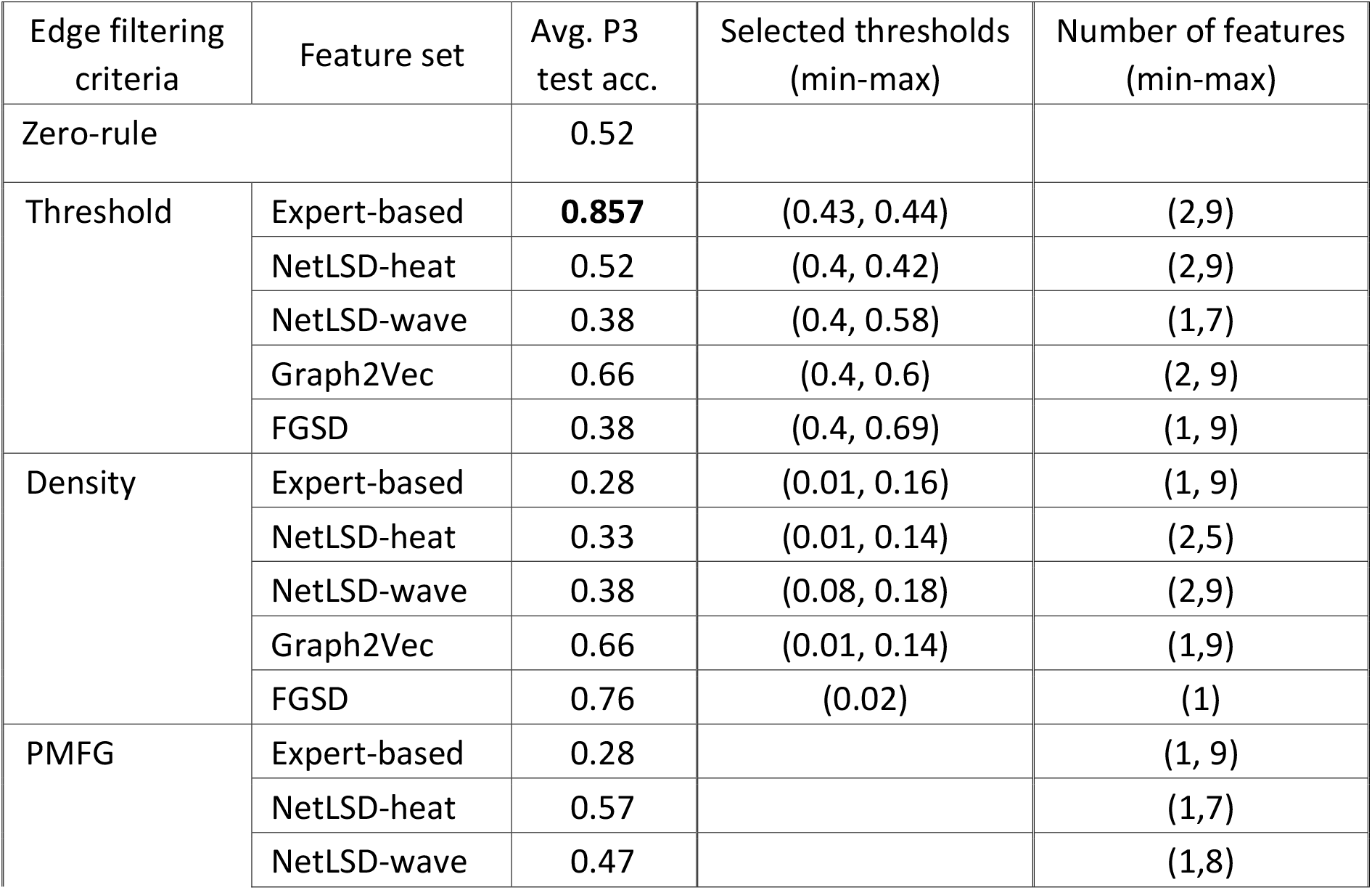
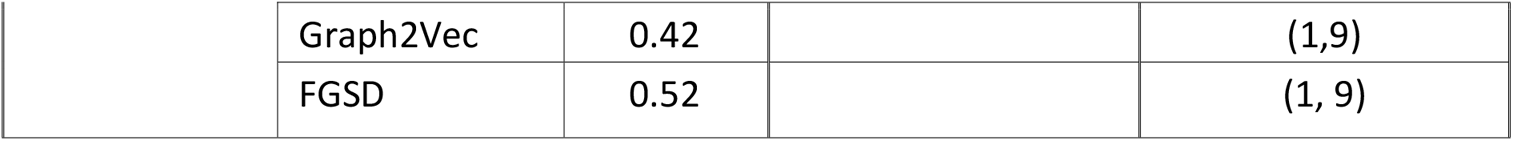
Results of the tasks with the different edge criteria, feature sets and number of features for predicting high/low CBM after rehabilitation treatment.

#### Sensitivity analysis

Models sensitive to small changes in hyperparameters may not be reliable and may not generalize well to new data. Sensitivity analysis reported here is important especially due to the small size of the analyzed dataset. To assess how sensitive the Δ*CBM* prediction pipeline to changes in the hyperparameter values is, we measured the average change in accuracy as a function of the hyperparameter values. The analysis relies on results from the inner (validation) loop of NLOOCV.

##### FGSD is too sensitive to density fluctuations

We analyzed the sensitivity of Δ*CBM* prediction based on FGSD with one dimension and the density edge filtering criteria. Figure 3.a presents the validation accuracy as a function of the density. The accuracy spikes to 0.76 at a density of 0.02. Despite high accuracy, the model performance shows high sensitivity to density fluctuations. Around a density of 0.02 ± 0.001, the accuracy score drops down to 0.50-0.55.

**Figure 3:**
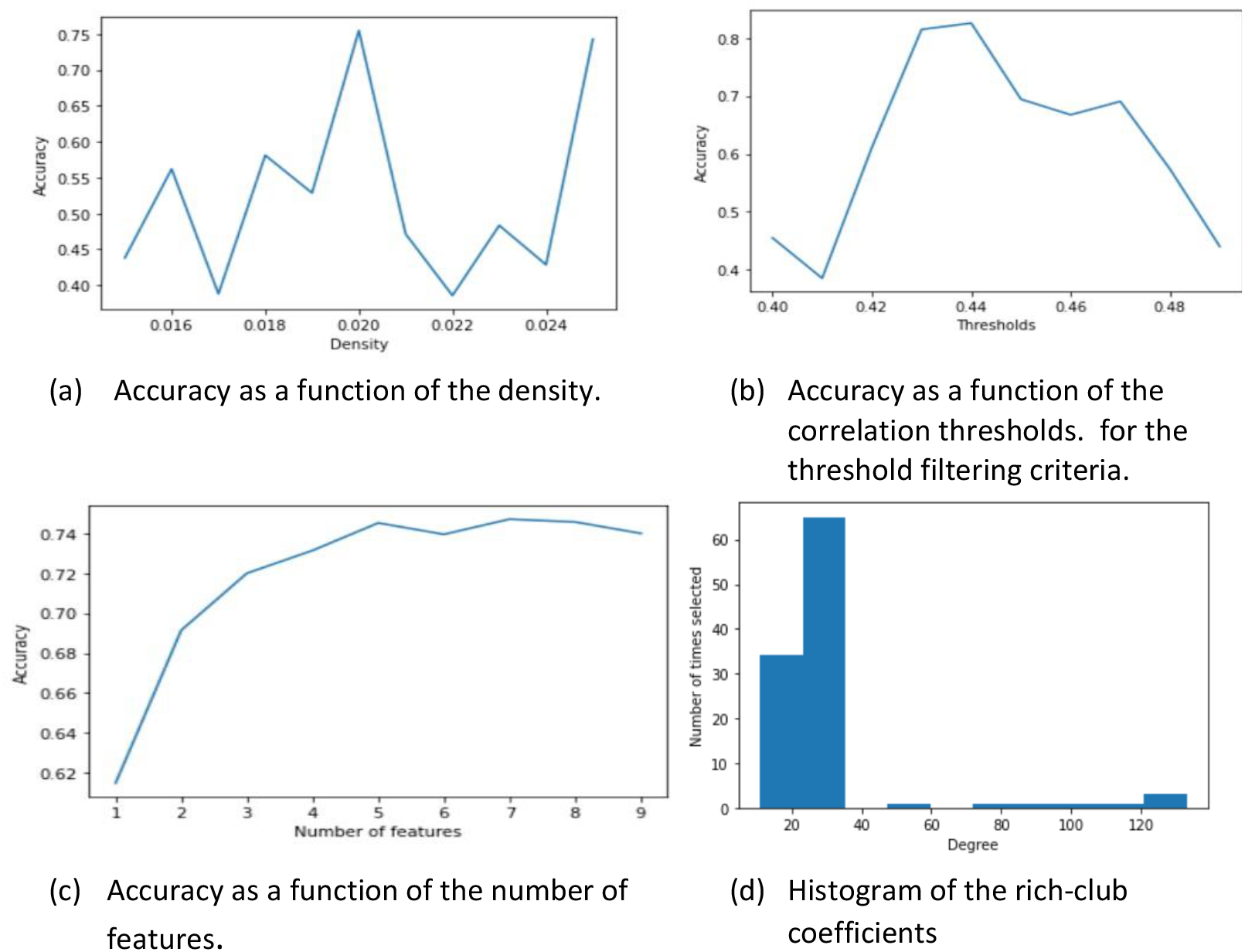
Parameter sensitivity results. (a), (b), and (c) display the average accuracy as a function of density, correlation threshold (t), and number of features (k), respectively. (d) displays the selection of rich-club coefficients. Results in (a) are displayed for FGSD with one dimension. Results in (c) and (d) are averaged over configurations with good correlation thresholds (t ∈ [0.43, 0.47]). Results in (b) are averaged over configurations with sufficiently large number of features (k ∈ [3, 9]).

##### More than three expert features are sufficient to produce good results

Figure 3.c presents the average validation accuracy of Δ*CBM* prediction as a function of the number of selected global features with the threshold edge filtering criteria. Like Figure 3.c, the results are averaged over 21 test iterations and all thresholds in the range *t* ∈ [0.43, 0.47]. The results indicate that the use of more than three features is sufficient to produce *accuracy* above 0.67.

##### Expert features are not sensitive to correlation thresholds around 0.43-0.47

Next, we analyze the sensitivity of Δ*CBM* prediction based on global features with the threshold edge filtering criteria. Figure 3.b presents the validation accuracy of Δ*CBM* prediction as a function of the edge filtering threshold with 3-9 expert-based features. The validation accuracy is averaged over 21 test iterations and seven feature set sizes. The accuracy as a function of threshold is bimodal, with the major mode around 0.45, and the minor mode around 0.47. For threshold values *t*, between 0.43-0.47, the accuracy is consistently above 0.7, with the best performance obtained for *t* ∈ [0.43, 0.44].

#### Consistency of the feature selection

Replicability and robustness are major concerns given the large number of features and limited dataset size. We therefore quantified the consistency of the hyperparameters and their optimization methods. A robust feature is one that performs consistently well for different brains and hyperparameter values.

To quantify the consistency of features, we used the NLOOCV process. As a result, in every test iteration (i.e., the external NLOOCV loop) different values of the hyperparameters were chosen. We report the most frequently chosen features and hyperparameter values and quantify their consistency in Table 3, where we show the number of times a feature was selected out of the possible iterations. This experiment is performed with a fixed number of features (k=6) and with six different thresholds (*t* ∈ [0.43, 0.47]), using 21 test iterations, (6 × 21 = 126). We consider multiple groups of the rich-club coefficients (1-9), (10,19), (20-29), (120,129). For each group we report the average number of times a feature from the group was selected out of 126 trials. Table 3 also can be viewed as a slice of Figure 3.c, where the number of features is six.

**Table 3:** The most consistent features. The number of times a feature was selected out of 126 iterations with threshold t ∈ [0.43, 0.47], six features, and all the test-sets (21).

| Feature name | Consistency: number of times selected (out of 126) |
| --- | --- |
| Rich-club coefficient (20-29) | 34 |
| Rich-club coefficient (10-19) | 21 |
| PageRank variance | 33 |
| Average neighbor degree of the left precuneus | 14 |
| Number of connected components | 11 |
| Average neighbor degree of the right inferior temporal cortex | 8 |
| Rich-club coefficient (0-9) | 5 |

Rich-club coefficients 10 to 29, Page rank variance, and average neighbor degree the left precuneus were found to be consistently informative for predicting CBM recovery. Figure 3.d shows the histogram of the rich-club coefficient. The x axis is the degree of the rich-club coefficient. The y axis is the total times this feature is selected. Rich-club coefficients 10-29 are selected most of the time as informative features. Higher rich-club coefficients are occasionally selected but are not reproduced through most of the experiment interactions, thus we do not consider them as consistent informative features. PageRank variance (local aggregated feature) is selected in 25% of the iterations. Additional meso-scale feature that was found to be consistently informative is the average degree of left precuneus neighbors. All neighbors in the functional graph were determined according to the selected correlation filtering thresholds (*t* ∈ [0.43, 0.47]).

### 3.3 Local features null result

When trained using unaggregated local features, all prediction models did not perform better than the zero-rule baseline. The feature selection process was also inconclusive as different sets of local features were selected in each iteration.

## 4 Discussion

### 4.1 Summary of the results

Our main research question is whether and how we can predict dynamic balance state and recovery based on resting state fMRI data. The results of the prediction of balance recovery following a rehabilitation treatment not only produced accurate (85.7% acc.) but also robust results, manifested by the repeated selection of hyperparameter values (e.g. the correlation thresholds) and features. The rich-club coefficients 10 to 29 were repeatedly selected as informative features for the prediction of CBM recovery, alongside PageRank variance, and average neighbor degrees of the left precuneus and the right inferior temporal cortex.

In addition, the single-dimension FGSD graph embedding performed surprisingly well with 76% accuracy. This result is sensitive to small changes in the density selection criteria, albeit consistently reproduced through variations in other hyper-parameters.

When predicting high/low dynamic balance state, analysis of T1 scans did not produce higher scores than the zero rule, whereas for T2 scans analyses resulted in accuracy level of 76% in retest and 86% in validation test when NetLSD embedding was used. The general inferiority of the state prediction models compared to the recovery models indicates that recovery mechanisms are more consistent across subjects than the underlying mechanisms of dynamic balance impairments after stroke.

### 4.2 Local and Global Features

Local features such as centrality measures or clustering coefficient describing specific ROIs did not perform well in either one of the three prediction tasks. We attribute the inferior predictive power of local features to the fact that the balance system involves several different regions of the brain (Takakusaki 2017). Thus, the balance system of stroke patients is not damaged in a specific ROI but rather the interconnection between the involved ROIs is malfunctioning (Almeida et al. 2017, Joubran et al. 2022). This hypothesis is partially confirmed by the average neighbor degree of the left precuneus and right inferior temporal cortex that showed a better predictive power than the degree measure (Table 3).

The most informative and robust features were the rich-club coefficient, Page rank variance, and average neighbor degree. These features had the highest information gain ratio and were selected repetitively in various settings. Rich-club coefficient describes how well a highly connected node is connected to other highly connected nodes. Ktena et al. (2019) showed in the past that rich-club coefficients can be used to leverage modeling post-stroke functional outcome. PageRank (Brin and Page 1998) is a measure of the importance of a node in a graph with random information flows. High variance of PageRank indicates that there are highly central nodes and highly peripheral nodes. Usually, centrality variance is the highest in sparse scale-free networks (Eguiluz et al. 2005) with few central nodes and many peripheral nodes.

Average neighbor degree is the average degree of the neighbors of a specific ROI. The average neighbor degree is a local aggregative feature (see Methods). The most selected average neighbor degree is of the left precuneus, which is a region that is related to episodic memory, spatial processing, and awareness. Wenderoth et al. (2005) Show that precuneus might be involved in shifting attention between different locations in space, which is necessary for dynamic balance control. The other region was the right inferior temporal cortex, a high-level area in the ventral vision stream, involved in perception and recognition (Baldauf and Desimone 2014).

### 4.3 Edge filtering

In our experiments the density edge filtering strategy did not exhibit favorable performance. PMFG criteria worked fine with 76% accuracy score in classifying the CBM score in the post-treatment (T2). The fixed threshold criteria performed similarly well with 76% accuracy score in classifying the CBM score in the post-treatment (T2) and 85.7% accuracy score in predicting the level of improvement in the CBM score at T2 relying on pre-treatment (T1) scan. The fixed threshold shows a better potential for addressing the proposed tasks and is recommended based on our results.

### 4.4 Embedding Algorithms

We used several algorithms to create an embedding of the graphs, while keeping the dimensions as low as possible due to dataset size. Some embedding algorithms rely on a ‘vocabulary’ of graphs to create an embedding to a graph. We conjectured that a small data-set of 21 subjects is not sufficient to supply such vocabulary. We used the algorithms as a black box without parameter-tuning except for the selection of the number of dimensions.

The algorithms we examined are NetLSD in waveform and heat form (Tsitsulin et al. 2018), FGSD spectral embedding (Verma and Zhang 2017), and graph2vec (Narayanan et al. 2017). Using algorithms for graph embedding or using pre-trained deep learning architecture can produce meaningful representations for the graphs. The most successful algorithm was NetLSD with 76% accuracy score in test-set in predicting the CBM score after treatment from T2 scan (P2). It also performed well with PMFG reaching 76% accuracy score for the same task. As for FGSD and graph2vec, both performed 76% accuracy score in predicting level of improvement in CBM from T1 scan (P3) and for predicting CBM score after treatment from T2 scan (P2) respectively. Both did not perform well in the sensitivity analysis and are therefore less recommended for this use case.

High performance of the graph embedding algorithms without expert-based features suggests the existence of undiscovered topological attributes with high predictive power for CBM recovery.

### 4.5 Nested Leave-One-Out Cross Validation

To both optimize the hyperparameters in a scientifically valid way and avoid overfitting we used additional nested LOO loops. Cawley and Talbot (2010) discuss the issues in cross-validation procedure when trying to optimize parameters. Due to the limited amount of data, it was important to use nested cross validation. The best practice for optimizing, evaluating, and testing, is using train validation and a test set. Training the model on a train set, optimize the hyperparameters by evaluating the performance on the validation set and test the model with the selected parameters and on the test set. In our case, we have only 21 subjects. We did not have sufficient subjects for splitting the data into train validation and test set. For this reason, we used the LOO. Since we also had to run a hyper-parameter optimization, we chose to use the nested LOOCV procedure.

### 4.6 Limitations

In this study we apply graph-theory driven tools to study a small dataset of graphs. The size of the dataset limits the kinds of analyses that can be performed and raises concerns of overfitting. A major limitation is that feature selection was based on information gain ratio, which measures only the importance of individual features and not the combined importance of sets of features. There might be better combinations of features that we are not aware of for the prediction tasks. Due to the small dataset, we could not have a highly dimensional feature space or use deep architectures but rather had to focus on robustness to avoid overfitting. Furthermore, to deal with the small dataset, we used LOOCV, which provided unstable results in some experiments, invalidating the respective models. Nevertheless, stable results in predicting CBM recovery are reassuring.

The proposed models predict the recovery of subjects that received rehabilitation treatment. However, since there was no control group that did not receive any treatment, we could not attribute the recovery only to the rehabilitation treatment. Other factors, such as the dynamic balance examination and passage of time could contribute to the recovery. Explaining the difference between the problems P1 and P2 is problematic as we are dealing with classification tasks. Even though we gained more robust results and higher accuracy scores in T2 than in T1, without analyzing the numeric change between the two groups we cannot make strong claims about differences between the two states. Larger datasets will allow us to address this question using a regression model.

### 4.7 Conclusions

Despite the limited data set, we report that global network connectivity features such as rich-club, embeddings, and average neighbor degrees can be utilized for predicting dynamic balance state and the outcome of a rehabilitation treatment after brain damage, even in small dataset. We show that nested cross-validation is a feasible approach for working with small datasets since it increases the reliability of the results. The local features that were extracted, such as average neighbor degree, were not related to motor control areas, raising the possibility that the informative dynamic balance markers are associated with sensory processing, and calls for additional research.

## Data Availability

All data produced in the present study are available upon reasonable request to the authors

## Ethics

This study was approved by the Research Ethics Committee of the Reuth Rehabilitation Medical Center, Tel-Aviv, Israel and by the Research Ethics Committee of Soroka University Medical Center, Beer Sheva, Israel. All participants signed an informed consent prior to undergoing assessments. The ClinicalTrials.gov identifier number is NCT02215590, 13/08/2014

## Data and code availability

Access to the raw clinical data is restricted according to the requirements set by the Research Ethics Committee of the Reuth Rehabilitation Medical Center, Tel-Aviv, Israel and by the Research Ethics Committee of Soroka University Medical Center, Beer Sheva, Israel.

The code developed for the analysis of the functional brain networks and the respective sanitized networks are available on GitHub https://github.com/cnai-lab/cbm-fmri.

## Author Contributions

**Or Symonitz:** Software, Investigation, Validation, Writing - Original Draft, **Katherin Joubran**: Data Curation, Investigation, Writing - Review & Editing, **Rami Puzis**: Conceptualization, Methodology, Supervision, Writing - Original Draft, **Lior Shmuelof**: Conceptualization, Data Curation, Writing - Original Draft, Supervision, Funding acquisition.

## 5 Supplementary Material

| Feature Type | Name |
| --- | --- |
| 12<br>Local features aggregated according to four moments (mean, variance, skewness, and kurtosis) | Degree |
|  | Eccentricity |
|  | Sub-graph centrality |
|  | Closeness centrality |
|  | Degree centrality |
|  | Eigenvector centrality |
|  | Clustering coefficient |
|  | Page rank |
|  | Harmonic centrality |
|  | Katz centrality |
|  | Load centrality |
|  | Triangles |
| 11 Global features | Number of connected components |
|  | Global efficiency |
|  | Number of isolates |
|  | Density |
|  | Average clustering |
|  | Degree assortativity coefficient |
|  | Degree Pearson correlation coefficient |
|  | Spectral bipartivity |
|  | Global reaching centrality |
|  | Edge connectivity |
|  | Diameter |
| Meso-scale features | Average neighbor degree |
|  | Mean degree connectivity |
|  | Rich-club coefficient |

